# Compassionate drug (mis)use during pandemics: lessons for COVID-19 from 2009

**DOI:** 10.1101/2020.05.07.20094839

**Authors:** Amanda M Rojek, Genevieve E Martin, Peter W Horby

## Abstract

**Background:** New emerging infections have no known treatment. Assessing potential drugs for safety and efficacy enables clinicians to make evidence-based treatment decisions, and contributes to overall outbreak control. However, it is difficult to launch clinical trials in the unpredictable environment of an outbreak. We conducted a bibliometric systematic review for the 2009 influenza pandemic to determine the speed, and quality of evidence generation for treatments. This informs approaches to high-quality evidence generation in this and future pandemics.

**Methods:** We searched PubMed for all clinical data (including clinical trial, observational and case series) describing treatment for patients with influenza A(H1N1)pdm09 and ClinicalTrials.gov for research that aimed to enrol patients with the disease.

**Findings:** 33869 treatment courses for patients hospitalised with A(H1N1)pdm09 were detailed in 160 publications. Most were retrospective observational studies or case series. 592 patients received treatment (or placebo) as participants in a registered interventional clinical trial with results publicly available. None of these registered trial results were available during the timeframe of the pandemic, and the median date of publication was 213 days after the Public Health Emergency of International Concern ended.

**Interpretation:** Patients were frequently treated for pandemic influenza with drugs not registered for this indication, but rarely under circumstances of high-quality data capture. The result was a reliance on use under compassionate circumstances, resulting in continued uncertainty regarding the potential benefits and harms of anti-viral treatment. Rapid scaling of clinical trials is critical for generating a quality evidence base during pandemics.

**Funding:** Wellcome Trust.

## Putting research into context

### Evidence before this study

Delays in conducting and disseminating high-quality research into potential interventions may limit pandemic responses. There is, however, little evidence to formally assess how research into therapeutics is conducted and published during a pandemic; this is needed to guide research policy design for pandemic preparedness going forward. During the 2003 Severe Acute Respiratory Syndrome (SARS) outbreak, limited data indicated a delay in publication of vaccine trials such that most results were published after the outbreak had ended. It has not been assessed if this was the case during the more recent A(H1N1)pdm09 pandemic, which was a much larger outbreak with broader geographic reach.

### Added value of this study

This study identifies that the vast majority of patients who were treated during the A(H1N1)pdm09 pandemic were given anti-virals outside of prospective interventional trials. It also quantifies a significant delay to publication of results of trials of these drugs, such that most were only available after the outbreak. These two factors limited the quality of evidence generated from treating a large number of individuals with these drugs. This is the only work to systematically assess how clinical evidence accrued during this pandemic.

### Implications of all the available evidence

These findings highlight a missed opportunity to generate high-quality clinical evidence during the A(H1N1)pdm09 pandemic. The testing of potential therapeutics during the current COVID-19 pandemic risks similar pitfalls; this study suggests that testing of novel therapeutics in this context should be as part of large multi-centre, prospective, controlled trials with rapid dissemination of results. Pandemic preparedness efforts must reduce the barriers to initiating and scaling such clinical research. A continued acceptance of treatment under a compassionate care paradigm fails patients and harms pandemic response.

## Introduction

Viral pandemics constitute a major threat to global health security. The potential for future influenza pandemics is considered likely. In the past 20 years, we have also seen the emergence of zoonotic human respiratory coronaviruses with pandemic potential. These have been Severe Acute Respiratory Virus (SARS; caused by SARS-CoV-1), Middle East Respiratory Syndrome (MERS) and, currently, COVID-19 (caused by SARS-CoV-2) which emerged in late 2019.^1^ Study of the 2009 H1N1 strain influenza A (A(H1N1)pdm09) pandemic, the largest respiratory viral outbreak in recent years, can provide insights into the research processes during a pandemic, with the aim of improving these for other outbreaks, including COVID-19.

One important element of pandemic mitigation is prophylaxis and treatment of patients. For emerging viral infections, antiviral therapies are a key medical countermeasure because vaccine production takes months or years, whereas effective antiviral medications may already exist. For COVID-19, the potential of several existing medications (including remdesivir, lopinavir/ritonavir and hydroxychloroquine and tocilizumab) is of interest. During the influenza A(H1N1)pdm09 pandemic, there was interest in neuraminidase inhibitors (NAIs) as anti-influenza agents, although adequate safety and efficacy data supporting their use were lacking. This evidence has now substantially strengthened^2^ however, much of this was generated after the pandemic. There has been no quantitative assessment of the volume and quality of information that is produced regarding treatments during the pandemic period. This data is important, because it represents what is available to clinicians making treatment decisions for patients under conditions of significant uncertainty,^3^ and during surging patient numbers.^4^

The objective of this systematic review is to investigate how safety and efficacy data for treatment of A(H1N1)pdm09 accrued during the pandemic (see table 1). We review the quantity and timing of publication of clinical trials of treatments, but also information where patients were treated outside a formal trial setting (case studies or series, and observational studies), or when research was registered but not completed, as these may represent both the best quality evidence available at the time, and also opportunities lost to gather high-quality evidence.

**Table 1:**
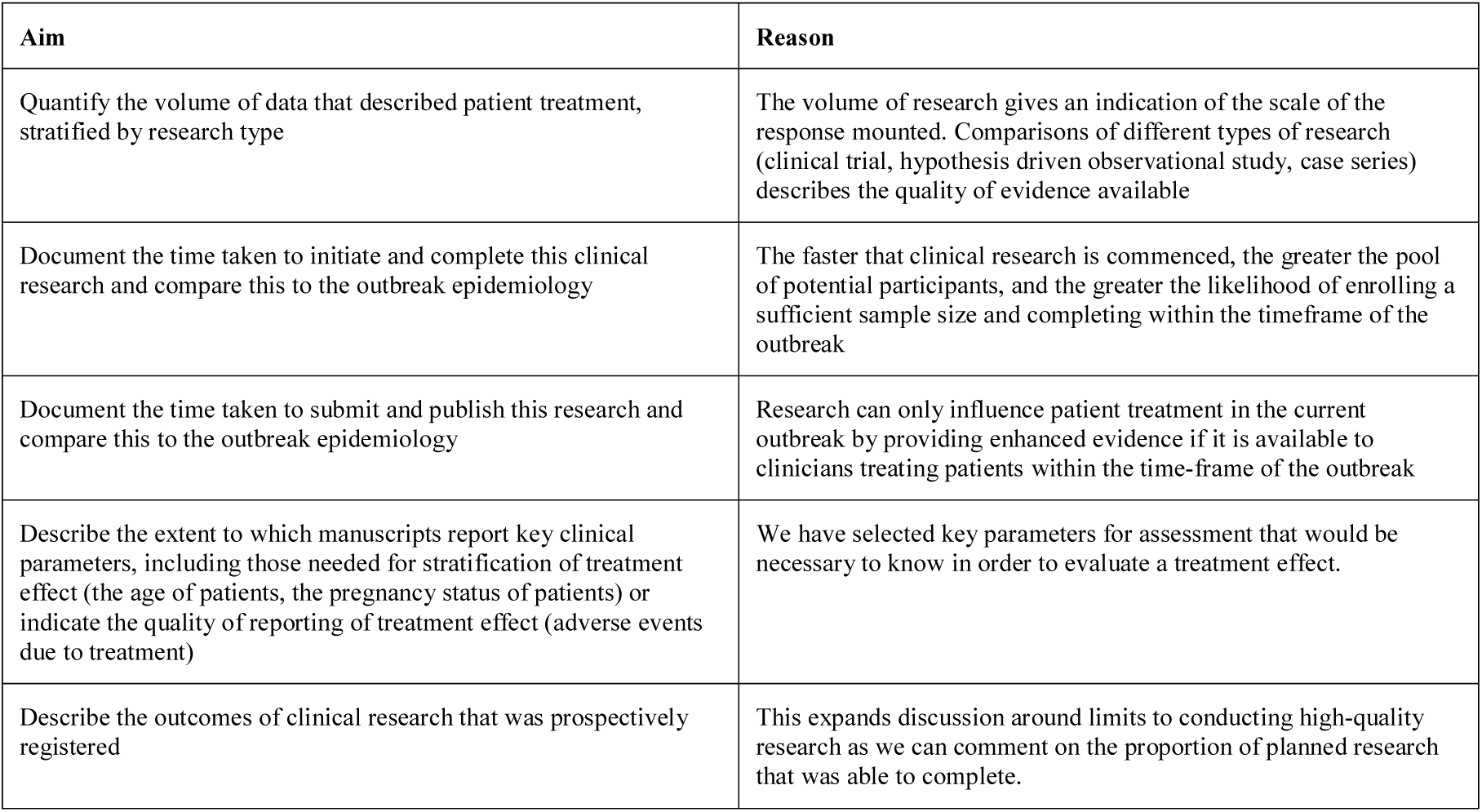
Detailed objectives and rationale with respect to evidence generation during a pandemic.

| Aim | Reason |
| --- | --- |
| Quantify the volume of data that described patient treatment, stratified by research type | The volume of research gives an indication of the scale of the response mounted. Comparisons of different types of research (clinical trial, hypothesis driven observational study, case series) describes the quality of evidence available |
| Document the time taken to initiate and complete this clinical research and compare this to the outbreak epidemiology | The faster that clinical research is commenced, the greater the pool of potential participants, and the greater the likelihood of enrolling a sufficient sample size and completing within the timeframe of the outbreak |
| Document the time taken to submit and publish this research and compare this to the outbreak epidemiology | Research can only influence patient treatment in the current outbreak by providing enhanced evidence if it is available to clinicians treating patients within the time-frame of the outbreak |
| Describe the extent to which manuscripts report key clinical parameters, including those needed for stratification of treatment effect (the age of patients, the pregnancy status of patients) or indicate the quality of reporting of treatment effect (adverse events due to treatment) | We have selected key parameters for assessment that would be necessary to know in order to evaluate a treatment effect. |
| Describe the outcomes of clinical research that was prospectively registered | This expands discussion around limits to conducting high-quality research as we can comment on the proportion of planned research that was able to complete. |

## Methods

We conducted a systematic search to identify patients treated for A(H1N1)pdm09 during the pandemic. We searched two types of evidence: peer-reviewed publications and clinical trial registration records. An experienced librarian advised on search strategy. We prospectively registered the review (PROSPERO database record CRD42016039549). Details of compliance to MOOSE and PRISMA guidelines are found in appendix 1.

### Published literature search

We searched the PubMed database according to the search strategy found in appendix 2. To capture information on how many patients received treatment outside of a trial, we included case studies, case series, and observational research in addition to interventional research. The single exception was to limit descriptions of the use of oseltamivir to publications with ten or more patients, because case reports were abundant. We included research that described hospitalised patients and reported acute clinical outcomes (defined as length of hospitalisation, intensive care admission or length of stay, medical complication, requirement for mechanical ventilation, or mortality). We included patients with only laboratory confirmed disease. While A(H1N1)pdm09 was the prevailing strain during the outbreak, inconsistencies in defining probable cases between papers meant a consistent inclusion method a was otherwise not possible. We included papers only if enrolment opened between April 1 2009 (when the virus strain was first identified) and was completed by August 10 2010 (the declaration of the end of the Public Health Emergency of International Concern [PHEIC]) by the World Health Organisation (WHO). This limitation was necessary to differentiate research conducted specifically for the pandemic, compared with routine seasonal influenza reporting once A(H1N1)pdm09 became a seasonal strain. These criteria did not apply for clinical trials (where there could be no confusion with seasonal reporting).

We excluded papers if description of treatment was not quantifiable, or the treatment name was absent (including use of the general term ‘antiviral therapy’). We defined treatment as pathogen-directed therapy (e.g. antivirals), or host-directed therapy where there was a specific indication for A(H1N1)pdm09. We therefore excluded descriptions of standard intensive care interventions including corticosteroids and extracorporeal membrane oxygenation. When a single patient cohort (same sample size, enrolment period, author(s), and study location) were presented in more than one paper, duplicates were excluded. We excluded languages other than English.

### Clinical trial registry search

We undertook two clinical trial registry searches. The purpose of the first was to examine research that was planned in response to the pandemic. ClinicalTrials.gov was searched using the condition ‘H1N1’ and dates were restricted to limit to registration dates following the onset of the pandemic. A second search was conducted to identify pre-existing influenza studies that were able to enrol A(H1N1)pdm09 infected patients. ClinicalTrials.gov was searched using the condition ‘influenza’. Detailed inclusion and exclusion criteria, and subgroup analysis plans for both searches are contained in appendix 3.

### Data extraction

One reviewer (AR) undertook data extraction according to the pre-specified inclusion and exclusion criteria. Decisions were recorded using electronic systematic review software (Rayyan^5^), available to the senior author (PWH). We did not request missing data from authors, as this does not contribute to the aims of this review. Details of the data extracted are in appendix 3.

### Statistical analysis

Descriptive statistics are presented as frequencies for categorical variables, and median with interquartile range for continuous variables. The findings from published literature and trial registries are reported separately. Analysis of the literature was stratified by research type. Chinese medicines are presented as a single class because individual components could not be differentiated. Assessment of combination therapy was not possible due to variable reporting practices in the literature. Stata MP/15.0 and Microsoft Excel for Mac/15.21.1 were used for statistical analysis and graphical depiction.

## Results

### Findings from published literature

This review includes 160 papers (summarised in figure 1, details in appendix 4) that describe 39577 hospitalised patients with A(H1N1)pdm09 and 33869 treatment courses (table 2). Twelve different treatments were used, with oseltamivir being most common (table 2). The median number of treatments described per manuscript is 63 (IQR 22-193). Of the 160 papers included, two are interventional trials (n=73, representing 0·2% of total reported patients),^6,7^ 28 are prospective observational studies (n=6102, accounting for 15·4% of total patients), 129 are retrospective observational studies or case reports (n=33342, 84·2% of total patients), and one enrolled patients both prospectively and retrospectively (n=98, 0·2% of patients).

**Figure 1:**
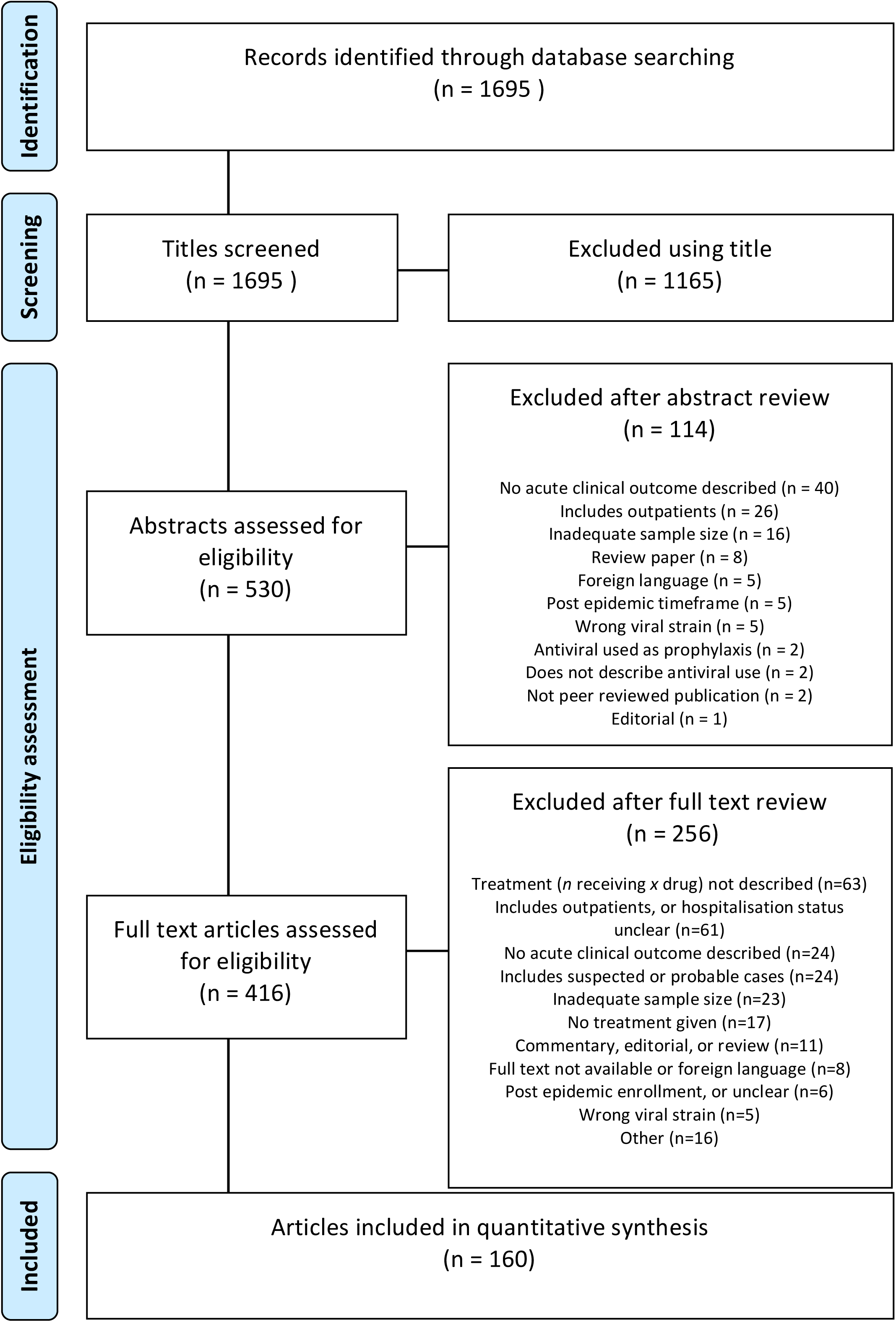
Study selection

**Table 2:** Volume of treatment courses described in the literature for hospitalised patients with 2009 H1N1 influenza during the pandemic period. * Some patients received more than one treatment. ** when publication describes use of that drug. ^where clear indication was influenza. eIND = emergency investigational new drug authorisation. EUA = emergency use authorization.

| Treatment name | Number of publications reporting use | Total number of patients receiving treatment* | Median (IQR) number of patients receiving treatment per publication** | FDA drug approval status for use in influenza in 2009 |
| --- | --- | --- | --- | --- |
| Oseltamivir | 154 | 31737 | 63 (21–188) | Approved for acute uncomplicated influenza, expanded under EUA April 2009 |
| Zanamivir | 54 | 368 | 2 (1–9) | Approved for acute uncomplicated influenza, expanded under EUA April 2009 |
| Peramivir | 14 | 403 | 1 (1–3) | Unapproved, eIND in April 2009, EUA October 2009 |
| Amantadine | 11 | 86 | 3 (1–13) | Approved for acute uncomplicated influenza, but resistance to A(H1N1)pdm09 demonstrated |
| Rimantadine | 5 | 32 | 3 (1–13) | Approved for acute uncomplicated influenza, but resistance to A(H1N1)pdm09 demonstrated |
| Ribavirin | 5 | 34 | 2 (1–6) | Not approved for influenza |
| Intravenous immunoglobulin | 4 | 44 | 4 (2–20) | Not approved for influenza |
| Chinese medicines | 3 | 1051 | 245 (56–750) | Not approved for influenza |
| Convalescent Plasma | 2 | 52 | 26 | Not approved for influenza |
| Macrolides^ | 1 | 31 | 31 | Not approved for influenza |
| Sirolimus | 1 | 19 | 19 | Not approved for influenza |
| Statins^ | 1 | 12 | 12 | Not approved for influenza |

Early initiation of prospective research maximises the probability of meeting sample size targets before an outbreak wanes. The median delay to first patient enrolment since identification of the pandemic viral strain (April 1, 2009) for prospective observational studies was 102 days (IQR 61-172 days). The two clinical trials began enrolment after a delay of 244 days and 275 days.

For prospective observational studies, enrolment stopped a median of 274 (IQR 195–313) days after viral identification. This was 223 days before the PHEIC ended (August 10, 2010), but when case numbers were falling. The two clinical trials closed enrolment 699 and 944 days after virus identification (March and November 2011).

The publication dates of all articles over time are shown in figure 2. No (0/2) interventional trials were published before the end of the PHEIC. 25% (7/28) of prospective observational studies, and 22% (28/130) of retrospective or mixed-enrolment research was published by the end of the PHEIC. The median date of publication for all papers was March 18, 2011 (IQR September 28, 2010–October 24, 2011); this was 213 days after the PHEIC ended. Overall the median delay between final patient enrolment (or inclusion) and date of publication of was 444 days (IQR 281–684). The median date between final patient enrolment and submission was 302 days (IQR 142–534), between submission and acceptance 93 days (IQR 63–144), and acceptance to publication was 56 days (IQR 24–94) where data existed for these intervals.

**Figure 2:**
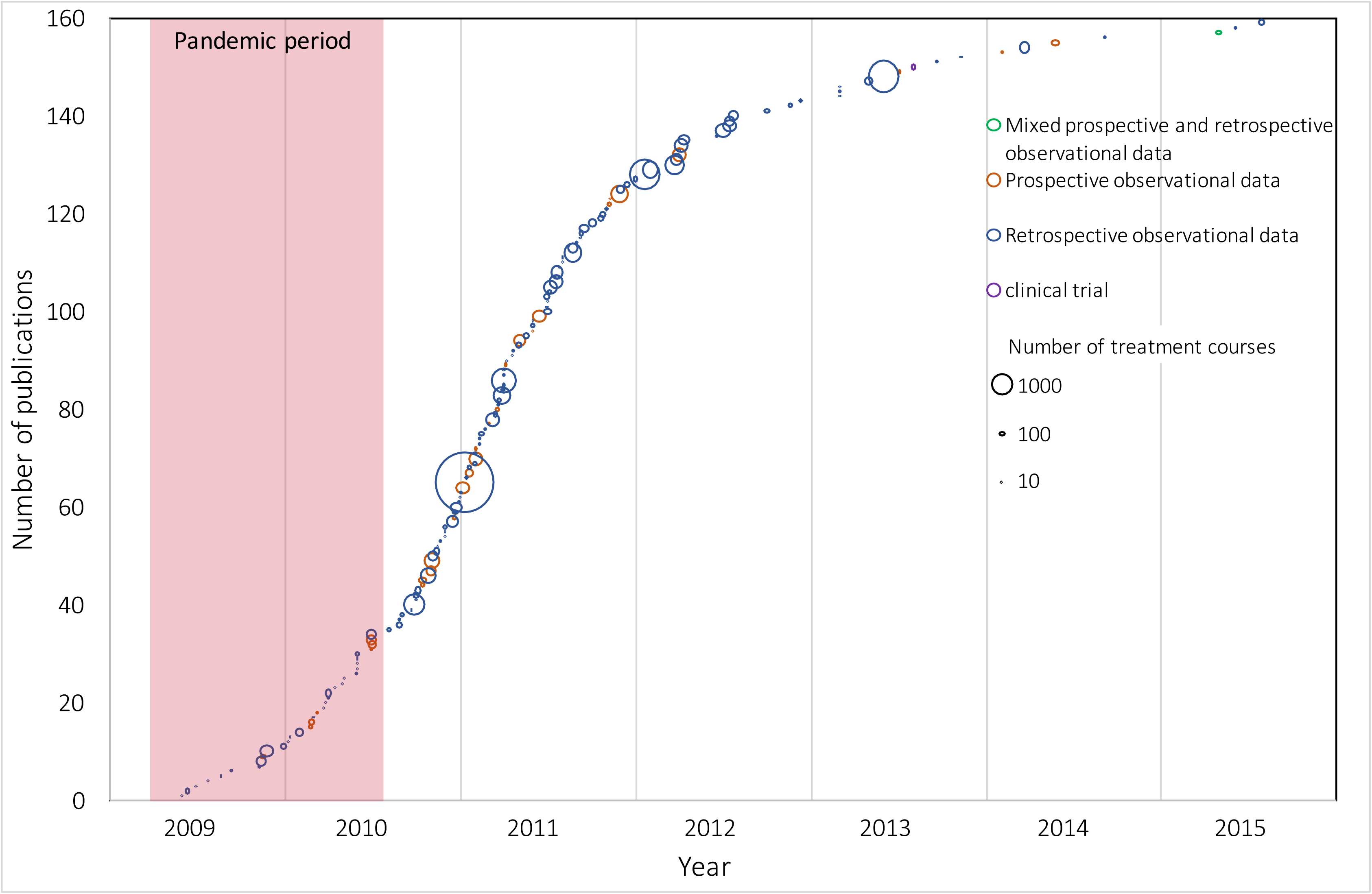
Publication of included studies over time, according to type of study and number of treatment courses described. The pandemic period ranges from the 1^st^ of April 2009, to the end of the PHEIC on 10^th^ of August, 2009.

39 countries reported treatment data. The highest number of papers were published by the United States (n=25, reporting 2559 treatment courses), followed by China (n=16, reporting 14680 treatment courses) and Spain (n=16, reporting 4103 treatment courses). Country level data describing the number of publications and treatment courses described, and the first date of patient enrolment in prospective research (where relevant) are shown in appendix 5).

Articles described the pregnancy status of patients in 88% (140/160) of articles. Articles described the inclusion of elderly patients in 88% (140/160) of articles and children in 93% (149/160) of articles.

23% (36/160) of papers described adverse effects from treatment had occurred. In 42% of cases, adverse effects or severe adverse events were noted. 13% (21/159) of articles tested for resistance and described some resistant samples, 4% (6/159) of articles tested for resistance but found no mutations, 3% (5/159) of articles reported clinical suspicion of antiviral resistance, and in the remaining 81% (129/159) of papers there was no statement regarding antiviral resistance; one paper described no antiviral use and was excluded.

### Findings from H1N1 trial registrations

Fifteen H1N1 study registration records were included in the review (appendix 6) comprising 10 interventional trials and 5 observational studies (2 with treatment efficacy outcomes, and 3 with general acute clinical outcomes) planned during the pandemic. A total of eight different treatments were to be studied; oseltamivir, zanamivir, convalescent plasma, intravenous immunoglobulin, rosuvastatin, sirolimus, Chinese herbs, and vitamin supplementation (vitamin A, C, E).

Of the 15 studies, nine are reported as completed, four were terminated due to the end of the H1N1 pandemic or declining case numbers, and the status of two studies is not recorded. The anticipated and actual enrolment of patients into all studies is depicted in figure 3. Some study protocols excluded patients because of young age (25%) and pregnancy (50%). Results are available in the literature for three of the completed studies (figure 3), representing 153 patients, and available on the clinical trials registry for an additional two of the terminated studies. A sub-group analysis of clinical trials that only included hospitalised or severe cases is provided in appendix 7.

**Figure 3:**
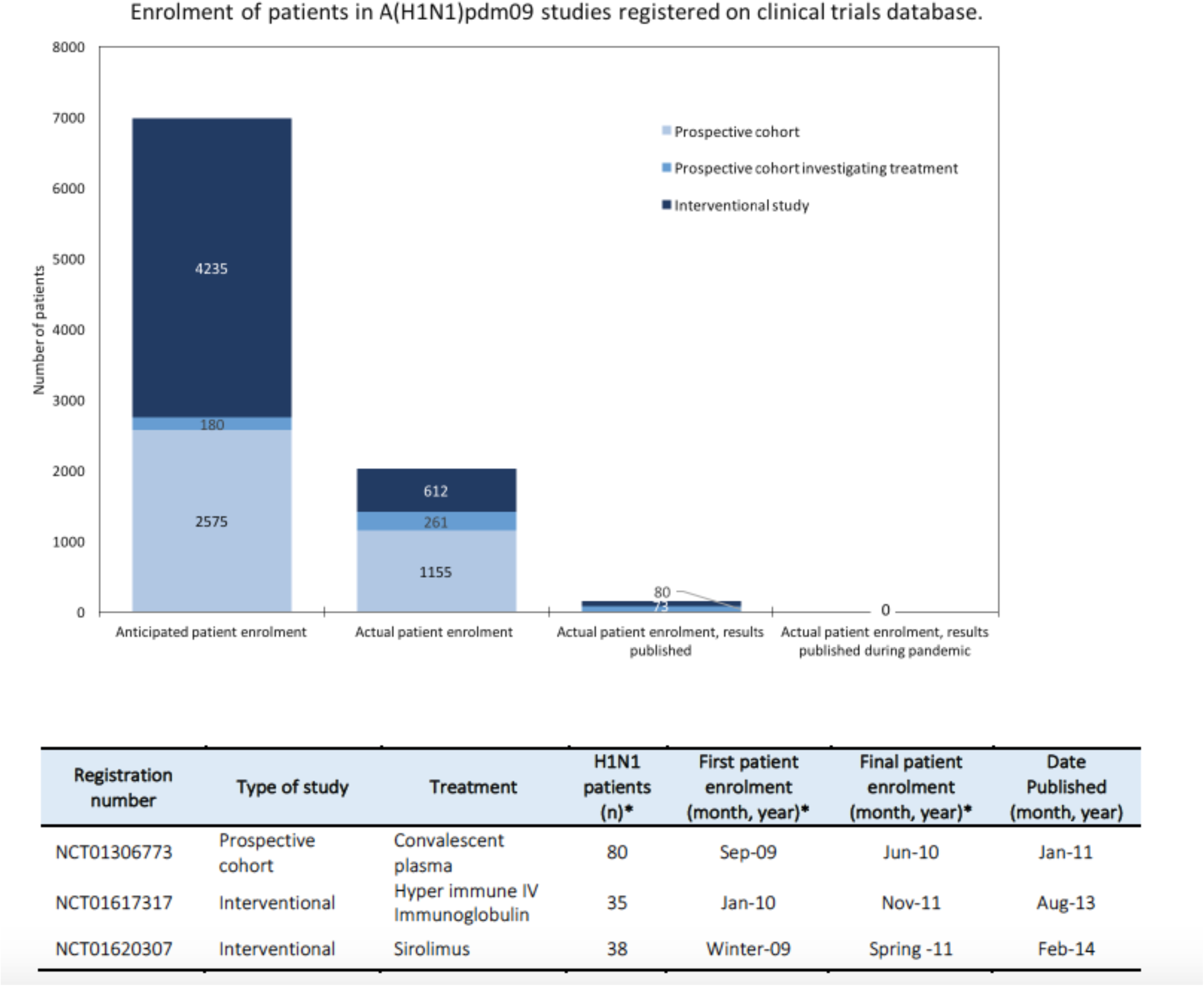
Anticipated and actual enrolment of patients in A(H1N1)pdm09 studies registered on clinical trials database. Table insert displays the enrolment number and publishing timeline for completed studies with results published. *where conflict existed between numbers in the clinical trial recod and publication, publication numbers were used.

### Findings from influenza trial registration

18 influenza registration records were reviewed (appendix 8). There were 16 interventional trials and two observational studies that were enrolling patients during the A(H1N1)pdm09 pandemic period. The treatments under investigation were oseltamivir, sambucol supplement, zanamivir, peramivir, amantadine, pomegranate supplement, nitazoxanide and favipiravir.

11 studies were completed, four were terminated early, the status of two is unknown, and one study has ongoing enrolment listed. Results were available for nine of 11 completed studies (table 3), representing 439 A(H1N1)pdm09 patients.

**Table 3:** Enrolment number and publishing timeline for completed studies where results are published in the literature (date followed by reference), or on the clinical trials registration site (date followed by Unp). *where conflict existed between numbers in the clinical trial record and publication, publication numbers were used. N/R not reported.

| Registration number | Type of study | Treatment | Planned enrolment (n) | Total patients* (n) | H1N1 patients* (n) | First patient enrolment* (month, year) | Last patient enrolment* (month, year) | Date Published (month, year) |
| --- | --- | --- | --- | --- | --- | --- | --- | --- |
| **Enrolment commenced before pandemic** |  |  |  |  |  |  |  |  |
| NCT00298233 | Phase 2 | Oseltamivir | 400 | 326 | 72 | Apr-07 | Feb-10 | May-13<sup>32</sup> |
| NCT00391768 | Phase 1/2 | Oseltamivir | 108 | 87 | 37 | Jan-07 | Apr-10 | Mar-13<sup>33</sup> |
| **Enrolment commenced during pandemic** |  |  |  |  |  |  |  |  |
| NCT00949533 | Phase 3 | Oseltamivir | 125 | 37 | unknown | Aug-09 | Oct-10 | Apr-16(Unp) |
| NCT00957996 | Phase 3 | Peramivir | 300 | 127 | 94 | Oct-09 | Oct-10 | Aug-13<sup>34</sup> |
| NCT01199744 | Prospective cohort | Zanamivir | N/R | 1575 | unknown | Nov-09 | Apr-10 | Mar-11(Unp) |
| NCT01014988 | Phase 2 | Zanamivir | 150 | 130 | 92 | Nov-09 | Sep-11 | Feb-14<sup>35</sup> |
| NCT01052961 | Phase 4 | Oseltamivir | 400 | 155 | 34 | Jan-10 | Jun-12 | Dec-13<sup>36</sup> |
| NCT01050257 | Phase 3 | Oseltamivir | 200 | 118 | unknown | Jan-10 | Sep-12 | Aug-13(Unp) |
| NCT01068912 | Phase 2 | Favipiravir | 384 | 530 | 110 | Feb-10 | May-12 | Feb-14(Unp) |

## Discussion

There is consistent criticism that the research response to disease outbreaks is fractured and delayed.^8-10^ There has, however, been little quantitative examination of these assumed insufficiencies. This paper demonstrates that despite over 33000 treatment courses being described for hospitalised patients with influenza A(H1N1)pdm09 during the pandemic, fewer than 600 received treatment (or placebo) as participants in a registered interventional clinical trial with results available in the peer-reviewed literature. None of these registered trial results were available during the timeframe of the pandemic, as were few of the findings from observational studies. This constitutes a significant failure to collect high-quality data.

Our findings demonstrate that we must make improvements in order to offer patients, and their treating clinicians, evidence-based care during pandemics, including the COVID-19 pandemic. Several drugs are being investigated as potential treatment for COVID-19, but we note that some early published studies have been poorly controlled.^11,12^ Based on scant scientific data, the Unites States Food and Drug Administration has approved the emergency use of hydroxychloroquine for the treatment of COVID-19 outside of clinical trials.^13^ It is imperative that we learn from the A(H1N1)pdm09 pandemic and ensure that trials of therapeutics are done under conditions which allow for the collection of high-quality, interpretable data to inform future clinical care.

We found that most descriptions of treatment courses were in retrospective observational studies or case series, with few prospective studies launched. There was relative success, however, in enrolling A(H1N1)pdm09 patients in ongoing or seasonal influenza studies – of the 582 patients enrolled in a trial, 439 were enrolled in this manner. This suggests that platform trials should be adopted as a way to expedite outbreak research. Using this approach, multiple treatments (or even multiple respiratory viruses) can be evaluated under an overarching protocol and regulatory framework, improving efficiency.^14^ Sleeper protocol research may also provide a solution. These pre-prepared and pre-approved protocols can lay dormant, waiting for cases of pandemic respiratory viruses, and allow prior assessment of the logistics and feasibility of the protocol. An example of this type of protocol exists in for severe acute respiratory infections (NCT02498587), and has been used to rapidly enrol patients with COVID-19.

Recommendations following the Ebola Virus Disease (EVD) epidemic in West Africa suggest that clinical research should be initiated, enacted and completed by the time an epidemic peaks.^15^ We found that from the time influenza A(H1N1)pdm09 was first detected, it was over three months before prospective data collection began, and eight months before the first interventional trial began recruitment. While these delays compare well to recent evaluations of delays for other epidemic observational research^16^ or clinical trials,^17^ it remains too slow.

Additionally, the small sample sizes of literature included in our review indicates a fractured research response. It has been estimated that a sample size of 800 patients is required to power a randomised controlled trial of an NAI in hospitalised patients.^18^ No prospective research identified here was that large. Beyond the benefits of increased enrolment and external validity, multicenter research has specific advantages in epidemics. It can compensate for unexpected variations in epidemiology at the regional level (such as the sudden end to the EVD outbreak in Liberia that prematurely halted a clinical treatment trial)^19^ or the temporary closure of health care facilities with nosocomial transmission (such as occurred during the Severe Acute Respiratory Syndrome outbreak of 2003).^20^

We report long delays between clinical data capture and publication in the peer-reviewed literature. This is consistent with analyses for other disease outbreaks, including epidemiology reporting for SARS (where only 7% of articles were published within the timeframe of the epidemic)^21^ and randomised controlled trials of pandemic H1N1 vaccines (where only 29% of clinical trial results were published almost a year following the end of the pandemic).^22^ The present WHO standard for interventional clinical trials is that main findings are to be submitted for publication within 12 months of study completion^23^ and although no analogous guidelines exist for observational clinical data, there are analogous scientific and ethical imperatives for timely reporting.

While the ramifications of delayed reporting are described for other fields^24^ there are specific imperatives for rapid data reporting during epidemics. For example, observational data must be accrued to design interventional trials (such as approximating the type and rate of outcomes). Emerging evidence can also prioritise trials so that the most promising continue recruitment when there are a declining number of cases late during an outbreak.^25^

Initiatives to minimise publication delay now include fast-track review for manuscripts likely to change clinical practice.^26^ Pre-approval of trial protocols and results-free review (where initial review excludes results and some discussion) are models which may assist in timely reporting. There is also support for pre-publication online release of preliminary findings. Indeed, the increasing utilisation of pre-publication servers (such as medRxiv) has been reflected in the COVID-19 outbreak. Dissemination of research findings via pre-publication (prior to peer-review) reduces delays, but carries risks for validation of methodology, accuracy of data, and interpretation of findings. The extraordinary number of COVID-19 articles being submitted to pre-publication servers^28^ has led to a rapid, open, peer-review platform for COVID-19 preprints,^29^ in an attempt to improve quality control.

The scope of our review was narrowed due to the high volume of clinical literature discussing A(H1N1)pdm09. In particular, we focused only on hospitalised patients where most antivirals were used.^30^ We included several publication types, including case series or observational studies to provide an estimate of patients who may have been eligible for inclusion in a clinical trial (noting this estimate does not represent the true number of hospitalised patients who were treated). The precision of any estimate is affected by excluding papers where treatment was not clearly defined and when pandemic strain influenza was not laboratory confirmed. Our estimates of patients enrolled in clinical trials is almost certainly an underestimation – much of the momentum toward compulsory registration of clinical trials^31^ occurred subsequent to the pandemic and trials may have been registered elsewhere and so it is likely that other trials were planned, initiated, or even completed without public knowledge. We restricted our observational data collection to that captured before the end of the PHEIC, and while we recognise that research continued to occur during the second and third waves of the epidemic, differentiating this work from routine seasonal influenza research became difficult. Only English manuscripts were reviewed, and there was only one reviewer for pragmatic reasons.

Here, we demonstrate how tolerance of treatment under compassionate care circumstances during the influenza A(H1N1)pdm09 pandemic was not matched with a commitment to capture high-quality data on treatment use and therefore failed the standards expected of modern evidence-based medicine. Moreover, we show that the data that was collected on patients was incompletely reported and published after prolonged delay. We recommend early initiation of multi-center collaborative trials, and pre-approved or sleeper protocols as potential solutions to improve accumulation of treatment data during a pandemic.

## Data Availability

Data will be available following correspondence with the authors.

## Contributors

AR and PH conceived and designed the study, had full access to all data in the study. AR and GM and drafted the manuscript with input from PH. AR carried out the statistical analysis, takes responsibility for the integrity of the data and the accuracy of the data analysis and is guarantor.

## Ethical approval

Ethics approval was not required for this work.

## Data sharing

Data are available from the lead author on reasonable request.

